# Field-of-view confounding shapes genetic discovery from self-supervised cardiac-imaging phenotypes

**DOI:** 10.64898/2026.09.01.26361959

**Authors:** Devansh Pandey, Vagheesh M. Narasimhan

## Abstract

Self-supervised models increasingly convert medical images into quantitative phenotypes for biological discovery, but statistical reproducibility does not establish that a learned phenotype represents the intended anatomy. We trained a video masked-autoencoder on 69,932 UK Biobank cardiac cine-MRI studies and performed genome-wide association analysis of its latent representation. Although 18 of 20 leading axes were heritable with well-calibrated statistics, the representation encoded substantial field-of-view information: body size, stature and imaging centre (linear-probe *R*^2^ = 0.55 for site); standard genomic-control and LD-score diagnostics did not identify this source of phenotype-level confounding. Restricting the field of view to the heart and residualising body and acquisition covariates *before* dimensionality reduction substantially attenuated linear and non-linear nuisance information while retaining cardiac signal. Adjusting the same covariates only during association testing attenuated nuisance associations but recovered substantially less of the cardiac-associated genetic signal, consistent with nuisance variation having already influenced the principal-component basis. The corrected representation identified new associated loci beyond those detected using supervised phenotypes at matched sample size, which shared genetic architecture selectively with cardiac-conduction traits and were localised to cardiac structures within the imaged field of view. Confounding in learned medical phenotypes can arise upstream of association testing, highlighting the importance of auditing and, where appropriate, correcting learned representations before association testing.

## 1 Introduction

Genome-wide association studies (GWAS) of imaging-derived phenotypes have mapped the genetic architecture of organ structure and function at scale, but they inherit a bottleneck: the phenotype is only as rich as the expert measurement that defines it^1,2^. Left-ventricular volumes, ejection fraction, and wall thickness capture a deliberately low-dimensional summary of the beating heart and, by construction, discard morphological and dynamic variation that a segmentation pipeline was never built to quantify. Self-supervised representation learning offers an appealing alternative: a model trained only to reconstruct masked image content learns a compact latent code without human labels, and that code can be scanned for genetic associations directly ^3,4^. Recent work has applied this idea to retinal fundus images^5^, brain MRI ^6^, and spirograms and photoplethysmograms^7^, in each case reporting loci beyond those found from expert features.

Self-supervised representations offer a richer alternative to expert-defined imaging phenotypes, but they encode all image content, not only the organ of interest. In cardiac cine-MRI this includes surrounding anatomy, body habitus and acquisition-specific features. If such nuisance variation is heritable or correlated with population structure, a GWAS of the learned representation can recover the genetics of the nuisance while conventional association diagnostics remain well calibrated. Whether this occurs, how it can be detected, and where in the analysis pipeline it must be corrected remain unresolved.

Here we use cardiac cine-MRI in 69,932 UK Biobank participants^8,9^ to ask what biological variation a self-supervised imaging representation captures when used as a quantitative phenotype. We find that a representation can be highly heritable and statistically well calibrated while its genetic architecture is strongly influenced by anatomical and acquisition-related factors outside the intended organ. We then examine how this nuisance variation affects dimensionality reduction, and compare association-stage covariate adjustment with correction of the representation before PCA. A two-stage approach combining cardiac field-of-view restriction with representation-level residualisation reduces measured nuisance information and yields reproducible cardiac-associated genetic signals that complement expert cardiac-MRI measurements and retain prospective clinical information. Together, these results illustrate a form of phenotype-level confounding that may not be apparent from conventional GWAS diagnostics, and provide one approach for detecting and mitigating it.

## 2 Results

### 2.1 A heritable learned cardiac phenotype can encode unintended biological variation

We trained a video masked-autoencoder (Video-MAE ^3^) on long-axis cine-MRI (2-, 3-, and 4-chamber views) from 69,932 UK Biobank participants, yielding a 2,304-dimensional per-participant embedding, and reduced it to 20 orthogonal principal axes for association testing. As a positive control, a GWAS of expert-derived left-ventricular ejection fraction in the same cohort recovered the established cardiomyopathy locus *BAG3* /*HSPB7* with well-calibrated statistics (genomic inflation *λ*_GC_ = 1.05; Supplementary Fig. S1), confirming expected behaviour of the genotype–phenotype association pipeline. Applying REGENIE ^10^ to the learned axes and estimating SNP-heritability with LD-score regression ^11^, 18 of 20 axes were significantly heritable (heritability *z >* 3) with well-behaved intercepts. These results initially suggested that the learned representation captured broadly heritable variation.

Examining the phenotypic and genetic correlates of these axes provided a different interpretation (Fig. 1). The leading axis was genetically correlated with body-mass index (*r_g_*= −0.735, *p* ≈ 10^−89^) and body-fat percentage (*r_g_*= −0.67), and its top locus was *FTO*, an established adiposity locus, not a cardiac locus. A second axis mapped to *WNT16* /*LRP5*, a stature/bone locus. The cine field of view is a wide thoracic slab in which the heart occupies only ∼25–35% of the frame and the spine is visible in the 2-chamber view, so the representation has ample opportunity to encode body habitus. Linear probing quantified this additional information: from the embedding alone we could decode imaging centre (*R*^2^ = 0.55), the scanner site, a proxy for geographically structured population substructure, alongside body size and stature, while cardiac ejection fraction was only weakly decodable (*R*^2^ = 0.37; per-axis *r* as low as 0.10). A representation that encodes acquisition site more strongly than it predicts the heart’s pump function cannot, without further auditing, be assumed to constitute a cardiac-specific phenotype.

**Figure 1:**
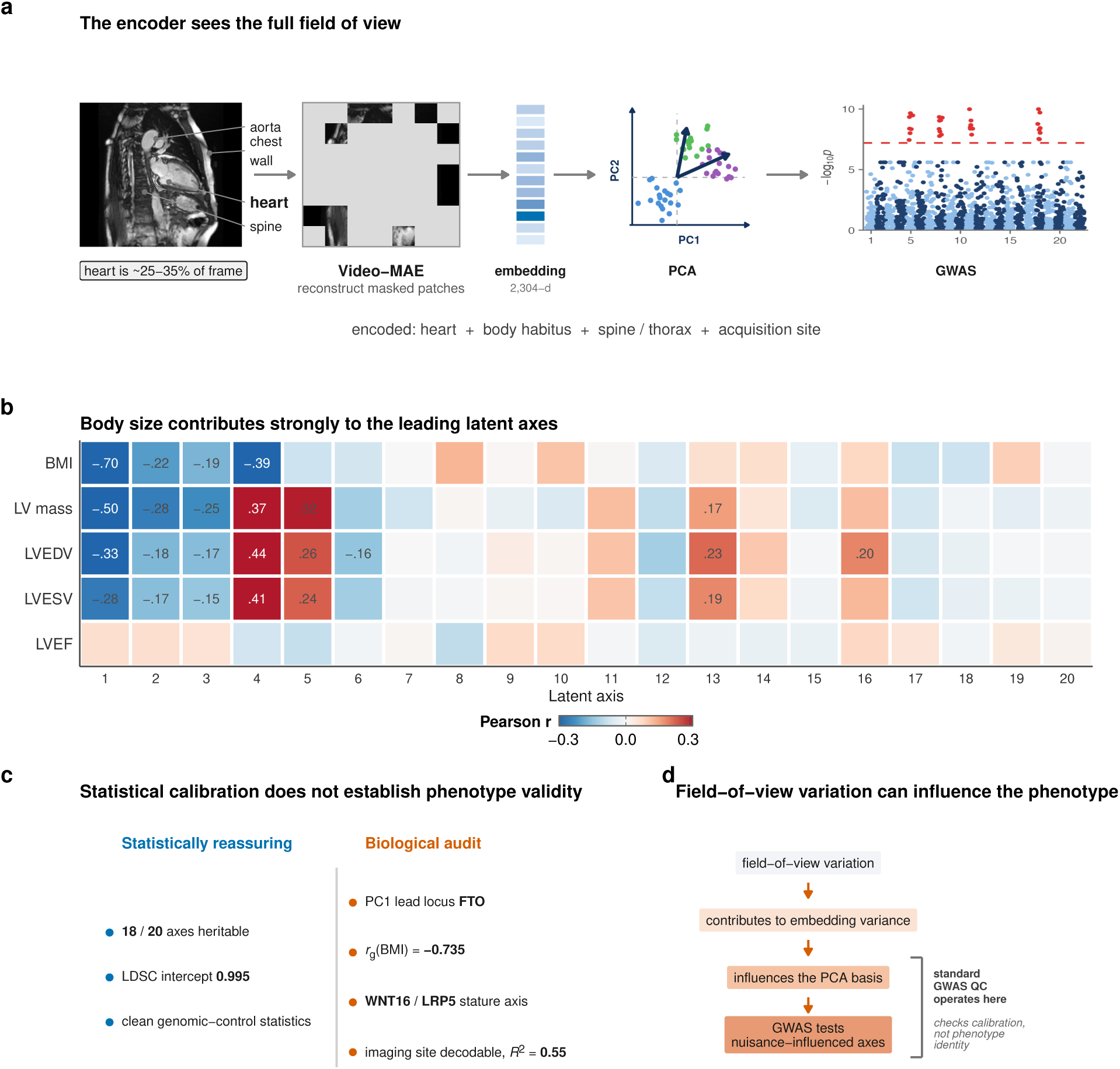
A heritable and well-calibrated learned cardiac phenotype can encode unintended biological variation. **(a)** A representative UK Biobank 2-chamber long-axis cine frame with labelled anatomy (aorta, chest wall, heart, spine); the heart occupies only ∼25–35% of the field of view, so body size, stature, spine and acquisition site are all available to the encoder, which is trained to reconstruct masked spatio-temporal patches of the whole image (cine MRI → Video-MAE → 2,304-dimensional embedding → PCA → GWAS). **(b)** Pearson correlation between each of the 20 raw latent axes and reference phenotypes; body size and left-ventricular mass load on the leading axes (BMI–PC1 *r* = −0.70) while ejection fraction is weak throughout. **(c)** The representation nonetheless passes conventional checks (18 of 20 axes heritable, LD-score intercept 0.995, well-behaved genomic control) even as a biological audit shows the leading axes track non-cardiac factors (PC1 top locus *FTO*; genetic correlation −0.735 with BMI; a *WNT16* /*LRP5* stature axis; imaging site decodable at *R*^2^ = 0.55). **(d)** Field-of-view variation contributes to embedding variance and can thereby influence the principal-component axes, so a GWAS of those axes tests nuisance-influenced directions, while standard GWAS quality control interrogates the association statistic rather than phenotype identity. The expert-LVEF positive control is shown in Supplementary Fig. S1.

Site decodability and heritability are not necessarily in conflict, because each latent axis can contain multiple sources of variation. Across the 20 raw axes, SNP-heritability showed a positive association with anthropometric loading (maximum |*r*| with BMI or height; Pearson *r* = 0.44, *p* = 0.053), whereas no association was observed with imaging-centre loading (*η*^2^ of centre; *r* = −0.09, *p* = 0.71; Supplementary Fig. S3). The two axes with the largest anthropometric loadings were also among the most heritable (PC1, |*r*|_BMI_ = 0.62, *h*^2^ = 0.167; PC4, |*r*|_height_ = 0.51, *h*^2^ = 0.160). Imaging-centre information was distributed across the representation: although centre was decodable from the full embedding (*R*^2^ = 0.55), it explained at most *η*^2^ = 0.09 of any individual tested axis. These observations are consistent with anthropometric variation contributing to the heritability of the raw representation while site contributes additional phenotypic variation.

Conventional GWAS quality-control metrics did not identify this feature of the phenotype. One axis with an LD-score intercept of 0.995, indicating well-calibrated association statistics, nonetheless showed a predominantly skeletal/anthropometric genetic signature, with lead signals at *WNT16* /*LRP5*. An intercept near one is consistent with little residual inflation from population stratification in the association statistics, but it does not establish that the phenotype itself represents the intended anatomy. Strong heritability and well-calibrated genomic control therefore do not establish that a learned phenotype measures the intended anatomy.

### 2.2 Nuisance correction before dimensionality reduction preserves additional cardiac-associated signal

We next asked at which stage of the analysis nuisance variation is best addressed, and compared correction of the representation before compression with adjustment at the association stage (Fig. 2).

**Figure 2:**
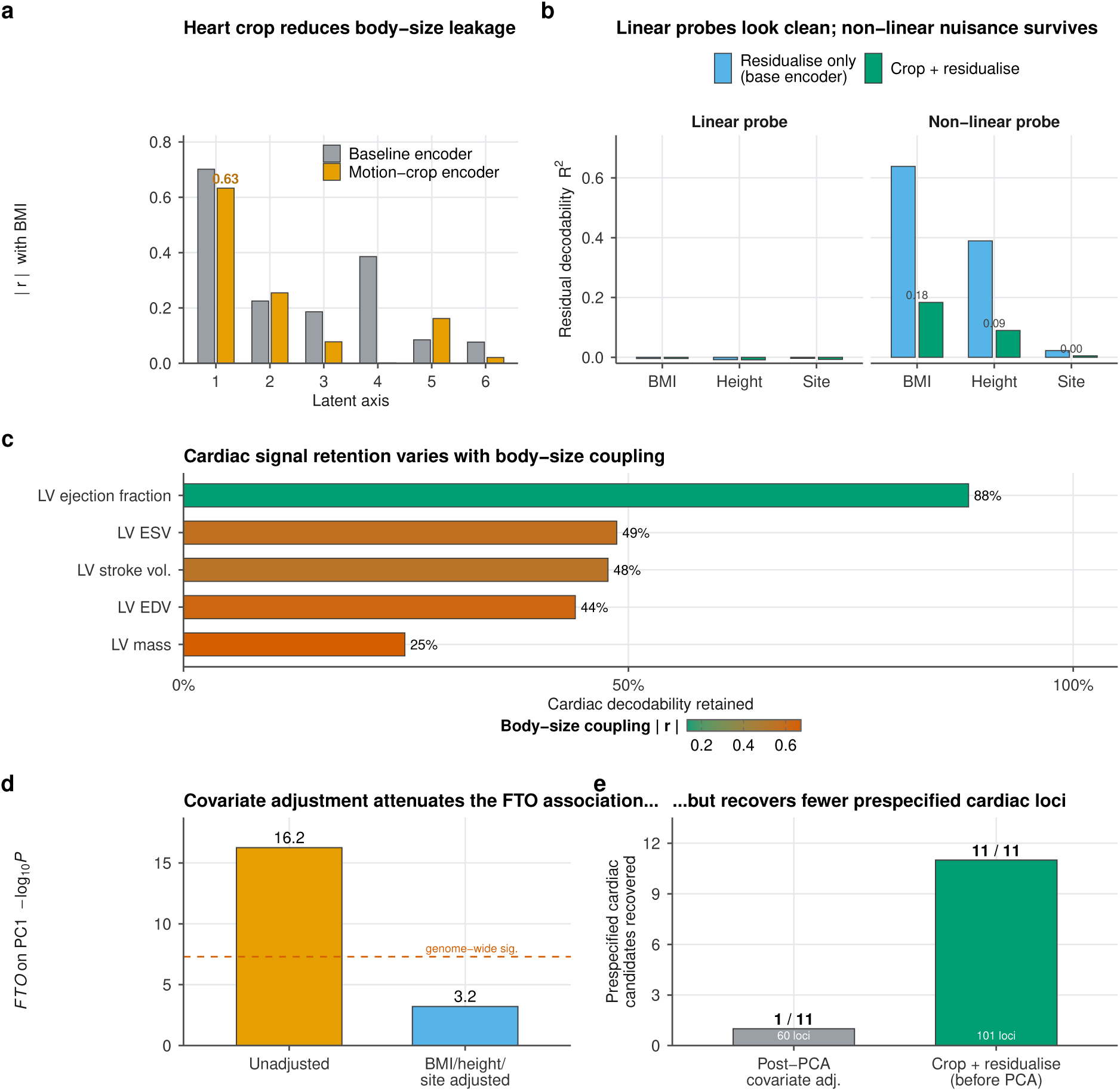
Representation-level correction before dimensionality reduction retains additional cardiac-associated signal. **(a)** Absolute BMI correlation on the leading axes for the baseline versus motion-crop encoder; cropping reduces but does not remove body-size leakage (|*r*| = 0.63 on PC1). **(b)** Residual nuisance decodability under linear and non-linear probes, for residualise-only (base encoder) versus crop-then-residualise: a near-zero linear probe leaves substantial non-linear body-size information (BMI *R*^2^ = 0.64) that crop-plus-residualisation substantially attenuates (BMI 0.18, height 0.09, site 0.005). **(c)** Cardiac decodability retained after correction, coloured by each phenotype’s body-size coupling: ejection fraction is spared (88%), body-size-coupled mass is not (25%). **(d)** Adding BMI, height and imaging centre as GWAS covariates collapses the *FTO* association on PC1 (log_10_ *P* 16.2 → 3.2). **(e)** Covariate adjustment recovers 1 of 11 prespecified cardiac positional candidates (60 loci) versus 11/11 for crop-then-residualise (101 loci), consistent with nuisance variance influencing the principal-axis basis.

We first retrained the encoder on a motion-derived cardiac crop, localising the beating heart from the per-pixel temporal standard deviation of the cine and removing ∼65–75% of the thoracic field of view, and then residualised the 2,304-dimensional cropped embedding on age, age^2^, sex, BMI, height and imaging centre before PCA. Cropping reduced but did not eliminate body-size information (BMI remained linearly decodable at |*r*| ≈ 0.63 on the leading axis), whereas residualisation suppressed linear nuisance decodability; non-linear probing showed that residualisation alone left substantial BMI and height information (random-feature *R*^2^ = 0.64 and 0.39 on the uncropped embedding), which crop-plus-residualisation attenuated to 0.18, 0.09 and 0.005 (BMI, height, imaging centre). The two operations were therefore complementary.

The information attenuated by correction tracks the body-size coupling of the underlying cardiac phenotype. Ejection-fraction decodability is largely preserved (*R*^2^ 0.492 → 0.434, 88% retained), whereas body-size-coupled chamber volumes and mass attenuate more (25–49% retained); across cardiac phenotypes the fraction retained falls monotonically with the phenotype’s correlation to body size (Supplementary Fig. S2). The correction is thus deliberately conservative about body-size-linked cardiac variation rather than indiscriminately lossy. Cardiac size does scale with body size, so some body-size signal is expected even from a perfectly heart-restricted phenotype. The field-of-view crop separates the two routes: body-size information that survives the crop reflects cardiac allometry and is then removed by residualisation at a known, quantified cost, whereas the much larger body-size signal in the uncropped representation, and all imaging-site information, is extra-cardiac.

We next asked whether a similar result could be obtained by retaining the raw principal axes and adjusting for the same nuisance variables only at the association stage. Adding BMI, height and imaging centre as GWAS covariates substantially reduced the *FTO* association on the leading axis (log_10_ *P*, 16.3 to 3.2, below genome-wide significance). However, the adjusted 20-axis omnibus recovered 1 of 11 prespecified cardiac positional candidates (*GJA1*; 60 loci), compared with all 11 after crop-and-residualise correction (101 loci). This difference is consistent with nuisance variation influencing the principal-component basis before association testing: when nuisance variance contributes strongly to the leading axes, lower-variance cardiac directions may be represented less strongly among the retained components. Association-stage adjustment can account for nuisance effects within the selected axes, but does not reconstruct variation that was not retained during dimensionality reduction. In this setting, correcting the representation before PCA therefore retained more of the cardiac-associated genetic signal.

### 2.3 Cardiac-associated genetic signals are reproducible after correction

GWAS of the residualised axes (*N* = 53,436) identified cardiac-associated signals that differed from those observed in the raw representation (Fig. 3). An omnibus test summing *χ*^2^ across the 20 orthogonal axes yielded 101 independent genome-wide-significant loci. One axis, PC4, was flagged as residual-confounded by a prespecified phenotype-level criterion (its residual genetic correlation with body-fat percentage, *r_g_*= −0.24, *p <* 10^−3^, the largest of any axis) applied independently of the association results; excluding PC4-driven loci and the MHC leaves 84 confound-controlled cardiac-associated loci. The main findings were similar when PC4 was instead omitted from the omnibus calculation, yielding a concordant 81 loci in which the same cardiac-development and conduction candidates were recovered. The loci lie near multiple genes with established roles in cardiac development and conduction, which we report as positional candidates (*TBX5*, *GATA4*, *PITX2*, *MYH6*, *TCF21*, *CCDC141*, *ZFPM1*, *THSD4*, and the gap-junction locus near *GJA1*); these overlap loci reported by expert-based cardiac-MRI GWAS ^12^ while extending into conduction biology. To assess whether overlap between the encoder training and GWAS samples (75%) materially influenced the associations, we repeated the analysis in 13,717 participants held out of encoder training; lead-SNP effect estimates were highly concordant with the full analysis (*β*-concordance 0.96–0.996).

**Figure 3:**
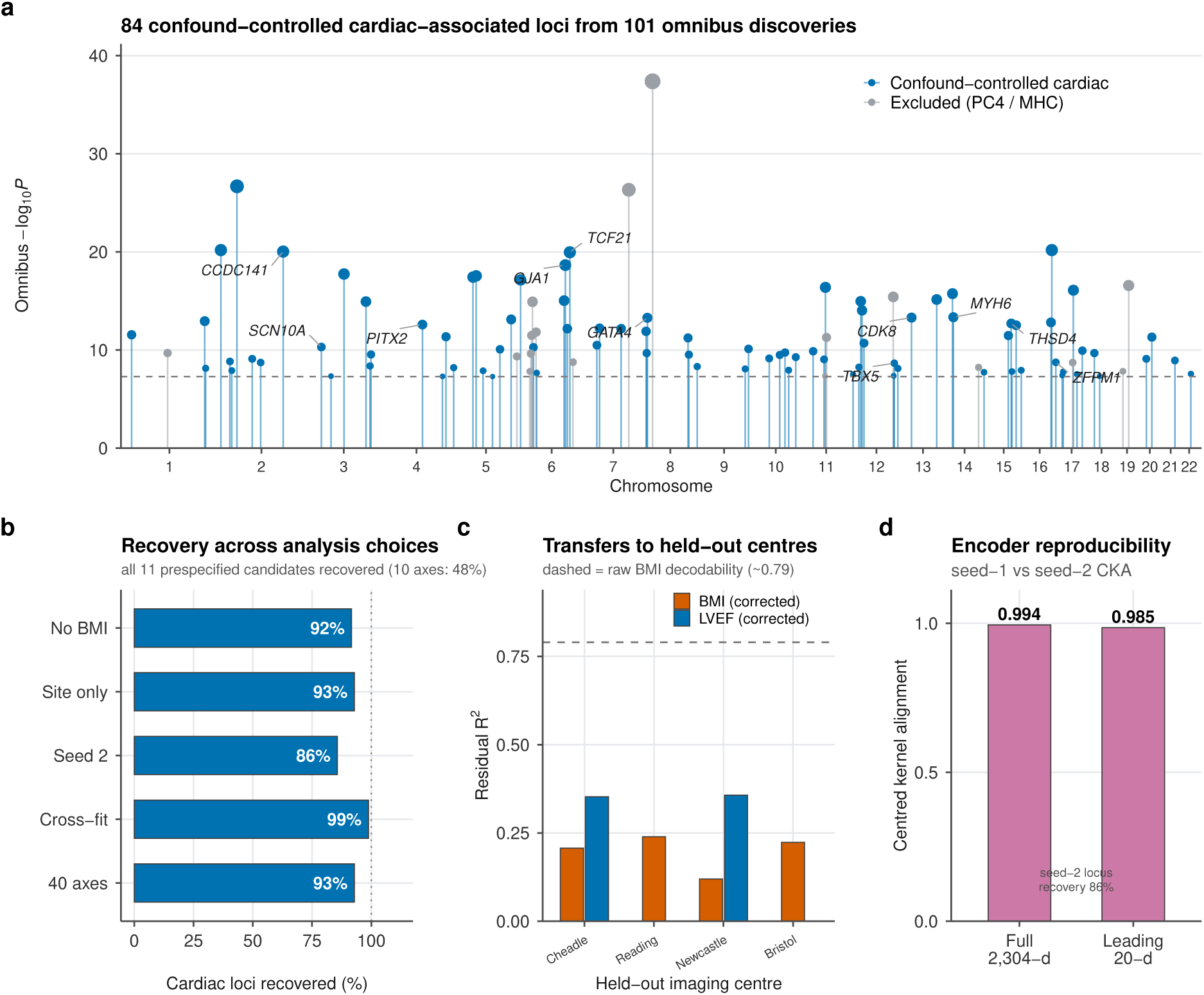
Cardiac-associated genetic signals are reproducible after correction. **(a)** Lead-locus plot of the confound-controlled omnibus GWAS (sum-of-*χ*^2^ across 20 axes): 101 independent genome-wide-significant loci, of which 84 comprise the confound-controlled cardiac set (blue; PC4-driven and MHC loci in grey). Labelled positional candidates are genes with established roles in cardiac development and conduction. **(b)** Cardiac loci recovered (lead SNP within 1 Mb of the main set) when the discovery is re-run without BMI, with imaging centre only, with a second encoder seed, with cross-fitted residualisation, or with 40 axes; every prespecified candidate is recovered each time (ten axes recover only 48%). **(c)** Leave-one-imaging-centre-out generalisation: residual decodability of BMI and ejection fraction in the held-out centre (dashed line, raw BMI decodability ≈ 0.79); corrected BMI falls to 0.12–0.24 while the ejection-fraction signal is retained, so nuisance attenuation transfers to acquisition centres not seen when the residualisation was fit. Ejection-fraction bars are shown only for Cheadle and Newcastle: the expert LVEF label (data field 22420) is largely unavailable for participants imaged at Reading and Bristol, so held-out LVEF decodability cannot be estimated there. **(d)** Encoder reproducibility: centred-kernel alignment between the seed-1 and seed-2 representations, overall and for the leading 20-dimensional subspace (seed-2 locus recovery 86%). Omnibus null calibration is shown in Supplementary Fig. S2.

The principal discoveries were broadly consistent across nuisance specification, cross-fitted residualisation, latent dimensionality and encoder retraining (Fig. 3): alternative nuisance models recovered 92–93% of the confound-controlled cardiac loci, five-fold cross-fitting (in which no participant’s phenotype is transformed with a model fitted on their own data) recovered 99%, 40 retained axes recovered 93%, and an independently trained encoder recovered 86%, with every prespecified candidate locus recovered in each analysis. Ten axes recovered only 48%, indicating that 20 axes provided a stable rather than minimal representation. At the representation level the two encoders were highly similar (linear centred-kernel alignment 0.99 overall, 0.985 between their leading 20-dimensional subspaces): although individual latent axes may rotate between runs, the leading representation subspace and locus-level associations showed high concordance. A leave-one-imaging-centre-out test showed that nuisance attenuation transfers to acquisition sites not seen when the residualisation was fit (held-out-centre BMI decodability 0.12–0.24 versus a raw ≈ 0.79, with ejection-fraction decodability retained; Fig. 3), and the omnibus statistic was well-calibrated (MOSTest between-axis *z*-correlations −0.08 to 0.05^13^; per-variant sum-of-*χ*^2^ bulk *λ*_bulk_ = 1.04 across 14.7 million variants; Supplementary Fig. S2). The corrected cardiac genetic signal is thus stable to the nuisance specification, the preprocessing strategy, the latent dimensionality, the acquisition centre, and independent encoder training.

### 2.4 Corrected representations contribute genetic signal beyond expert phenotypes

Having established a confound-controlled phenotype, we asked whether it contains genetic signal not captured by expert measurements (Fig. 4). Compared against a GWAS of 36 expert cardiac-MRI imaging-derived phenotypes, and controlling for the power advantage by down-sampling the learned-axis GWAS to matched sample size (≈33,000 vs 33,000), 19 of 33 independent loci (58%) had no genome-wide-significant expert-IDP association within the predefined locus/LD window; we refer to these operationally as learned-only loci. They include loci near the positional candidates *GJA1*, *PROX1* and *CDK8*. (In the full-power analysis this learned-only set extends further, e.g. to the conduction gene *SCN10A*.) The learned representation therefore contributes genetic signal beyond expert phenotyping, not a better-powered restatement of it. We present these genes as positional candidates: only two loci met the prespecified colocalisation threshold (PP.H4 *>* 0.5) against GTEx cis-eQTLs, both in aortic tissue, and, informatively, the locus we had labelled “*GJA1*” by proximity colocalised instead with a lncRNA, with *GJA1* itself 660 kb away, underscoring that proximity labelling is provisional. Nearest-gene annotation alone is therefore not evidence that the corrected phenotype measures cardiac biology, and we do not treat it as such; the two analyses below test cardiac specificity without relying on gene assignment.

**Figure 4:**
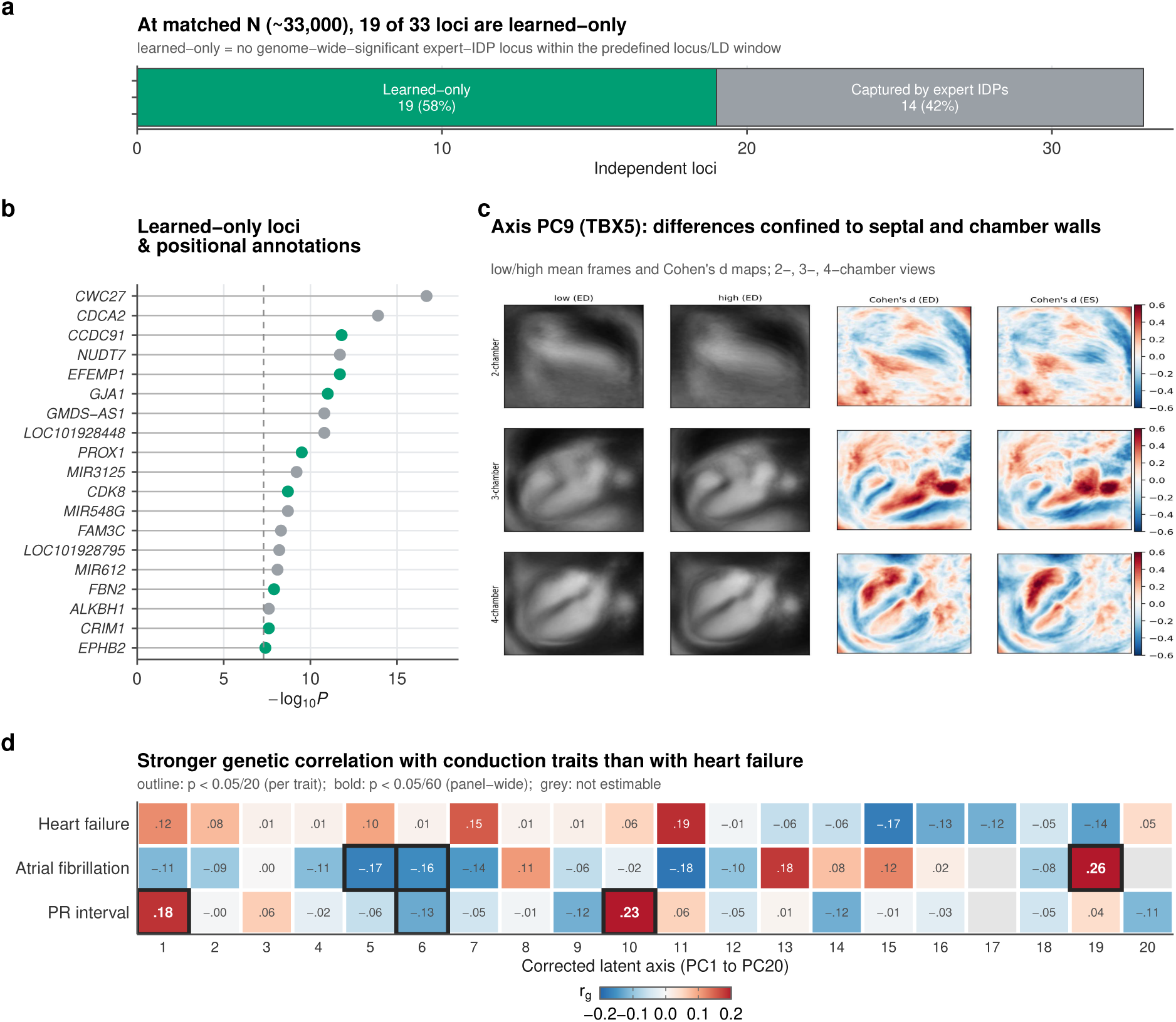
Corrected representations contribute genetic signal beyond expert phenotypes. **(a)** At sample size matched to a 36-trait expert cardiac-IDP GWAS, 19 of 33 independent loci (58%) are learned-only. **(b)** Ranked learned-only positional candidates (*GJA1*, *PROX1*, *CDK8* and others highlighted). **(c)** Cohen’s *d* effect-size maps for axis PC9 (positional candidate *TBX5*) across the 2-, 3- and 4-chamber views and end-diastolic and end-systolic phases, on the motion-cropped heart field of view: differences are confined to septal and chamber-wall structures rather than distributed across the frame. Maps are computed on cardiac images only and therefore show within-heart spatial structure, not organ-level specificity. **(d)** Genetic correlation of each corrected axis with the PR interval, atrial fibrillation and heart failure; two axes reach panel-wide significance for the PR interval and the PITX2/CCDC141 axis for atrial fibrillation, while heart failure is null (∗ *p <* 0.05*/*20; ∗ ∗ *p <* 0.05*/*60).

Three analyses that do not depend on proximity-based gene assignment provide additional evidence that the corrected axes are enriched for cardiac-related signal. First, the corrected loci carry excess association signal in independent cardiac GWAS. Testing the mean *χ*^2^ of the confound-controlled lead variants in external summary statistics against 10,000 size-matched random SNP sets, the loci were enriched for left-ventricular ejection fraction (8.1×, *p* = 1 × 10^−4^), PR interval (7.9×, *p* = 1 × 10^−3^), QT interval (5.3×, *p* = 1.8 × 10^−3^), atrial fibrillation (5.0×, *p* = 3 × 10^−3^) and heart failure (2.4×, *p* = 4 × 10^−4^) (Supplementary Fig. S4). Applying the same test to the loci from the *raw* axes, the enrichment profile differed substantially before and after correction: cardiac-trait enrichment was higher after correction (ejection fraction 2.4×→ 8.1×; atrial fibrillation 0.8×, non-significant, → 5.0×; QT 2.3×→ 5.3×), whereas skeletal enrichment was an order of magnitude lower (heel bone-mineral density 24.0× → 2.3×) and BMI enrichment was null after correction (1.0×, *p* = 0.50). Residual height enrichment persists after correction (5.2×), as expected from cardiac allometry. Second, the corrected axes are spatially structured within the heart rather than reflecting global image variation: Cohen’s *d* effect-size maps between the extremes of each axis, computed across the 2-, 3- and 4-chamber views and the end-diastolic and end-systolic phases, show differences confined to discrete cardiac structures rather than diffuse whole-frame shifts; for example, PC9 (positional candidate *TBX5*, a cardiac-development transcription factor) produces coordinated septal and chamber-wall differences consistent across all three imaging planes. Because these maps are computed only on cardiac images, they establish spatial structure within the heart but not organ-level specificity relative to other anatomy. Third, they share germline architecture selectively with cardiac *conduction* rather than disease: against a prespecified panel, two axes correlate with the electrocardiographic PR interval at panel-wide significance (PC10, *r_g_* = 0.23, *p* = 5 × 10^−6^; PC1, *r_g_* = 0.18, *p* = 7 × 10^−4^) and the PITX2/CCDC141 axis correlates with atrial fibrillation (PC5, *r_g_*= −0.17, *p* = 9 × 10^−4^), whereas heart failure shows no correlation surviving correction. This pattern is biologically consistent with the established roles of *PITX2* and other conduction-associated loci. Together, the enrichment for independent cardiac-trait association, the within-heart spatial structure and the selective conduction correlation provide evidence for cardiac-related signal that does not rely on gene labelling, although none of these establishes the specific biological mechanisms underlying individual loci.

### 2.5 Genetic architecture and clinical prediction capture distinct aspects of the learned phenotype

Finally, we asked how the corrected phenotype behaves as a clinical predictor, and found that genetic architecture and prospective prediction answer different questions (Fig. 5). In the imaged cohort we fitted Cox proportional-hazards models for incident atrial fibrillation, heart failure, coronary artery disease and cardiomyopathy (first-occurrence ICD-10 diagnoses after imaging, prevalent cases excluded), comparing age+sex, age+sex+expert-CMR measurements, and the latter plus the 20 corrected embedding axes. Adding the embedding improved five-fold cross-validated discrimination over expert CMR for atrial fibrillation (ΔC-index +0.024; *p* = 3 × 10^−24^; 571 events), heart failure (+0.021; *p* = 4 × 10^−4^; 205 events) and coronary artery disease (+0.011; *p* = 6 × 10^−7^; 756 events); cardiomyopathy was underpowered (53 events). The largest increment was observed for atrial fibrillation, consistent with the enrichment of associated loci near genes implicated in atrial and conduction biology (*PITX2*, *TBX5*, *GJA1*).

**Figure 5:**
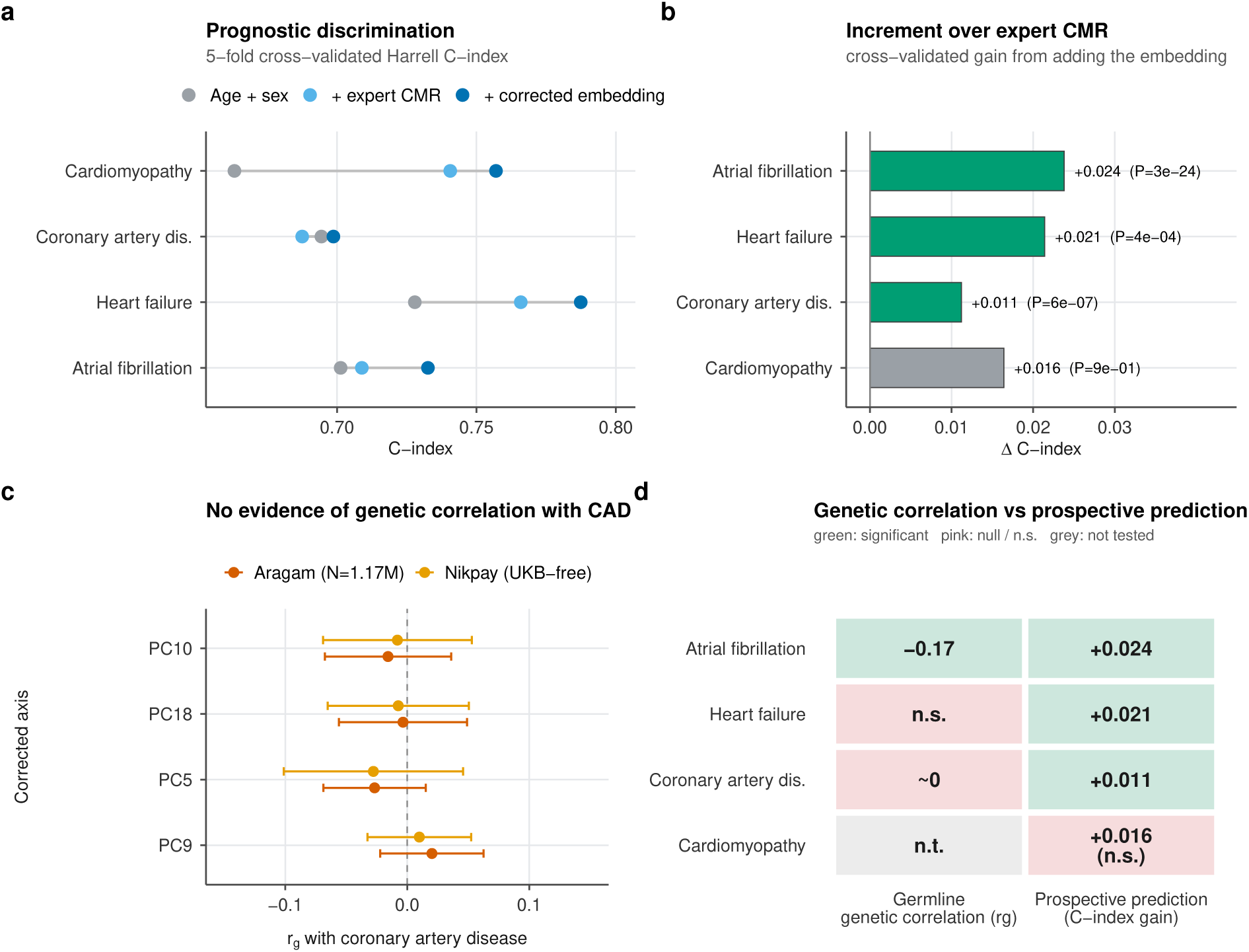
Genetic architecture and prospective prediction provide complementary views of the learned phenotype. **(a)** Five-fold cross-validated Cox C-index for age+sex, +expert-CMR, and +the 20 corrected embedding axes, for incident atrial fibrillation, heart failure, coronary artery disease and cardiomyopathy. **(b)** Cross-validated C-index increment from adding the embedding over expert CMR, with the full-data likelihood-ratio *p*; significant for AF, HF and CAD (strongest for AF), underpowered for CM. **(c)** No evidence of genetic correlation between the corrected axes and coronary artery disease was observed against either external GWAS (Aragam, *N* = 1.17M; Nikpay, UK-Biobank-free). **(d)** Genetic correlation versus prospective prediction by outcome (cells give the germline *r_g_* and the cross-validated C-index gain). CAD is the clarifying case: no evidence of germline genetic correlation but positive prediction, so inherited architecture and the acquired imaging phenotype are distinct.

Coronary artery disease provides an informative contrast between genetic correlation and prospective prediction. We found no evidence of genetic correlation with CAD in either of two well-powered external GWAS, Aragam et al. (*N* ≈ 1.17 million) ^14^ and Nikpay et al. (CARDIoGRAMplusC4D, UK-Biobank-free) ^15^: estimated *r_g_*values were close to zero and non-significant across axes. Yet the same representation improved CAD-event prediction. These results illustrate that germline genetic overlap and prospective phenotypic prediction need not coincide: a learned phenotype can be prognostically useful without sharing a disease’s inherited architecture. The learned cardiac phenotype showed little detectable overlap with CAD’s inherited genetic architecture yet carried prognostic information, a dissociation that the corrected representation makes explicit.

## 3 Discussion

A learned medical-image phenotype can be reproducibly heritable and show well-calibrated association statistics while still primarily encoding unintended biology. When nuisance content, here body size, stature and imaging site, is itself heritable and population-structured, a genetic analysis of the raw representation recovers the genetics of the nuisance while conventional diagnostics remain well-behaved. This represents a form of phenotype-level confounding that differs from the statistical confounding addressed by genomic-control and LD-score diagnostics: those tools ask whether the association *statistics* are confounded, not whether the heritable *phenotype* measures the intended organ. As learned phenotypes proliferate^5–7^, these results show that calibration of GWAS statistics should not be interpreted as evidence that the underlying learned phenotype is biologically specific.

Our results further suggest that the stage at which nuisance variation is addressed matters. Nuisance variation affects phenotype *construction*, not merely the subsequent association statistic: when it contributes strongly to embedding variance it influences the leading principal-component basis, reducing representation of lower-variance cardiac directions among the tested axes. Adjusting for the same covariates in the association model removes their effect from an already-selected axis (the *FTO* association collapses from log_10_ *P* = 16.3 to 3.2) but recovers only 1 of 11 cardiac candidates, against all 11 when the representation is corrected before compression. In this analysis, representation-level correction before dimensionality reduction retained substantially more cardiac-associated signal than association-stage adjustment. Similar considerations may apply to other analyses in which learned representations are compressed before association testing.

After correction, several complementary analyses were consistent with greater cardiac specificity: enrichment of the corrected loci for association in independent cardiac GWAS, matched-power learned-only loci, spatial structure confined to discrete cardiac structures within the imaged field of view, and selective genetic correlation with cardiac conduction (PR interval, atrial fibrillation via *PITX2*). We emphasise that the latter two lines of evidence, unlike nearest-gene annotation, do not presuppose which gene a locus acts through; establishing the causal genes and mechanisms at these loci will require functional follow-up. The correction itself has costs and limits. It can only attenuate confounding along measured, modelled covariates, and it deliberately removes body-size-coupled cardiac variation (ejection fraction retained at 88%, chamber mass at 25%); the recurrent residual-height genes (*ADAMTSL3*, *EFEMP1*) are a reminder that the correction is partial rather than absolute. The signal that survives this conservative correction and reproduces across nuisance specifications, cross-fitting and independent encoder training therefore has substantially stronger evidence of cardiac specificity.

Prediction and organ-specific genetic inference make different demands on the same representation, and coronary artery disease is the clarifying case. For prediction, field-of-view and acquisition-related features may contribute predictive information, whether through biological, demographic or site-associated pathways, and the corrected representation improved CAD-event discrimination despite showing no evidence of genetic correlation with CAD, consistent with the companion study’s finding that imaging risk is largely orthogonal to polygenic risk^16^. For organ-specific genetic inference, the same content can be severely misleading: a GWAS of the raw representation recovers the genetics of body size, not the heart. This study has limitations. Analyses are restricted to European-ancestry participants; a reserved multi-ancestry imaged sub-cohort awaits genotype access for replication, important because CAD genetic architecture differs across ancestries^17^. Locus-level discovery was stable across two independently trained encoders, but we did not establish one-to-one alignment of individual latent axes across training runs; the unit of inference is the locus, not the axis. Because the correction targets measured nuisance variables, unmeasured anatomical or acquisition-related confounding may remain. The interpretability maps were computed only on cardiac images; because no non-cardiac organ was tested, they demonstrate spatial structure within the heart but cannot establish that the axes are anatomically specific to the heart relative to other organs. Colocalisation used bulk adult tissue and is underpowered for developmentally acting transcription factors, the interpretability maps reflect morphology in static phases, and no external cine-MRI + genotype cohort of comparable scale currently exists for imaging-level replication. Our results support incorporating nuisance-decodability audits into analyses of learned imaging phenotypes, considering representation-level correction where appropriate, and comparing learned and expert phenotypes at matched statistical power before interpreting novel biological associations.

## 4 Methods

### 4.1 Cohort, genotyping and quality control

Analyses used cardiac cine-MRI and genome-wide genotype data from the UK Biobank^8^, a prospective cohort of ∼500,000 participants, under approved application 65439. Genotypes were imputed and quality-controlled by the central UK Biobank pipeline ^9^; we used the imputed release (GRCh38) and retained variants with imputation INFO *>* 0.8 and minor-allele frequency *>* 0.001 for association testing. Sample-level quality control removed participants with sex-chromosome aneuploidy, discordance between self-reported and genetic sex, outlying genotype missingness or heterozygosity, and consent withdrawals. Primary analyses were restricted to participants of genetically inferred European ancestry (projection onto 1000 Genomes principal components); one participant from each pair with a kinship coefficient corresponding to third-degree or closer relatedness was removed to yield an unrelated set. Ten genetic principal components were computed within the retained European sample and used as association covariates. Age was taken at the imaging visit, and body-mass index, standing height and sex from the imaging-visit assessment.

### 4.2 Cardiac imaging and preprocessing

Long-axis cine series (2-, 3- and 4-chamber views) were acquired on the standardised UK Biobank cardiovascular MRI protocol ^1^. After quality filtering, 69,932 participants’ cine series were processed. For each participant the three long-axis views were stacked as three input channels; frames spanning the cardiac cycle were resampled to a fixed temporal length and each frame resized to a fixed spatial resolution, with per-series intensity normalisation. Expert cardiac imaging-derived phenotypes (IDPs), including left-ventricular ejection fraction (data field 22420; *N* = 39,684 populated) and left-ventricular volumes and mass, were taken from the UK Biobank return of automated segmentation ^2^ and used as positive controls and as the benchmark for the incremental-value analysis. Preprocessing parameters are listed in Supplementary Table S1.

### 4.3 Self-supervised representation learning

We trained a video masked-autoencoder (Video-MAE ^3^; a vision-transformer encoder–decoder) on the stacked long-axis cine tensors. Training used tube masking of spatio-temporal patches with a mean-squared reconstruction loss computed on masked patches only; no phenotype, covariate or genotype information entered training. After pretraining, the decoder was discarded and the encoder token outputs were mean-pooled to a 2,304-dimensional per-participant embedding. The *base* encoder (uncropped field of view) is used for the confound diagnosis (Fig. 1); the *crop* encoder (below) underlies the discovery pipeline. To establish stability to the training run we independently pretrained a second base and a second crop encoder from a different random seed and repeated the entire downstream pipeline. Representation-level similarity between seeds was quantified by linear centred kernel alignment (CKA), computed on the full 2,304-dimensional embedding and on the leading 20-dimensional principal subspace. The network architecture and training schedule are given in Supplementary Table S1; a fixed random seed (42) was used unless otherwise stated, with the independent replicate trained from seed 202.

### 4.4 Motion-based cardiac field-of-view crop

The crop encoder restricts the field of view to the beating heart. For each participant we computed the per-pixel temporal standard deviation across the cine frames, which is largest over the contracting myocardium and blood pool; we thresholded this motion map at its 92nd percentile to localise the cardiac region, took the bounding box of the supra-threshold pixels between their 2nd and 98th percentiles, expanded it by a 12-pixel margin, and cropped every frame to this box (discarding ∼65–75% of the thoracic field of view; Supplementary Table S1). The Video-MAE encoder was then retrained from scratch on the cropped inputs with the same objective and schedule as the base encoder.

### 4.5 Representation-level residualisation

The crop-encoder embedding **E** ∈ R*^N^*^×2304^ was residualised column-wise by ordinary least squares on the design matrix **X** = [ **1**, age, age^2^, sex, BMI, height, imaging centre] (imaging centre entered as indicator variables), retaining the residual **R** = **E** − **X**(**X**^T^**X**)^−1^**X**^T^**E**. *Residualisation was applied to the embedding before dimensionality reduction*: principal components were computed on **R** (centred, unit-variance scaling), and the leading 20 axes were standardised to mean 0 variance 1 and carried forward as phenotypes. This ordering matters because nuisance variance that contributes strongly to **E** otherwise influences the principal-component basis itself (Fig. 2). As sensitivity analyses we retained 10 and 40 axes, and estimated the residualisation by five-fold cross-fitting (the OLS coefficients were fit on four folds and applied to the held-out fold, so no participant’s embedding was transformed with a model fitted on their own data).

### 4.6 Confound-decodability probes

To quantify how much nuisance and cardiac information a representation carries, we decoded each target (BMI, height, sex, imaging centre, and each expert cardiac phenotype) from the embedding with two probes, each evaluated by five-fold cross-validated *R*^2^ (balanced accuracy / AUROC for categorical targets). The *linear* probe was ridge regression; the *non-linear* probe was ridge regression on *D* = 1024 random Fourier features approximating a radial-basis-function kernel, included specifically to detect non-linear residual confounding that a linear probe cannot see. Probes were run on the base, crop, residualised-base and crop-then-residualised embeddings. Retention of a cardiac phenotype was defined as the ratio of its post-correction to pre-correction cross-validated *R*^2^. To assess whether the discovered loci carry cardiac-trait association independently of nearest-gene annotation, we compared the mean association *χ*^2^ of the lead variants in external GWAS summary statistics (LV ejection fraction, PR interval, QT interval, atrial fibrillation, heart failure; and height, BMI and heel bone-mineral density as non-cardiac references) against a null formed by 10,000 randomly drawn SNP sets of the same size from the same summary statistics, reporting the ratio of observed to mean null *χ*^2^ and the empirical permutation *p*. Random sets were matched on set size only and not on allele frequency, LD or local gene density; this comparison therefore quantifies empirical excess association relative to random variants rather than establishing functional enrichment, and the raw-versus-corrected contrast, in which both sets are lead variants of the same kind of analysis, is the more informative comparison. The same test was applied to the lead variants of the raw (uncorrected) axes, clumped to 1 Mb, to compare what the discovery captures before and after correction. To test whether axis heritability is carried by anthropometric rather than acquisition content, we regressed per-axis SNP-heritability (from the raw-axis LD-score regression logs) on two per-axis loadings computed in participants with complete covariates (*N* = 54,745): anthropometric loading, the maximum absolute Pearson correlation of the axis with BMI or standing height; and imaging-site loading, the proportion of axis variance explained by assessment centre (*η*^2^ from a one-way analysis of variance). Associations are reported as Pearson correlations across the 20 axes. Leave-one-imaging-centre-out generalisation was assessed by fitting the residualisation on all but one centre and evaluating decodability in the held-out centre.

### 4.7 Association testing

Per-axis genome-wide association was performed with REGENIE v4^10^. Step 1 fitted a whole-genome ridge-regression polygenic model on high-quality LD-pruned common array variants (MAF *>* 0.01, INFO *>* 0.9, pairwise *r*^2^ *<* 0.9 in 1 Mb windows) in blocks of 1000 variants, producing leave-one-chromosome-out (LOCO) predictions that control for polygenic background and cryptic relatedness. Step 2 tested imputed dosages (INFO *>* 0.8, MAF *>* 0.001) conditional on the LOCO predictor. Covariates were age, age^2^, sex, genotyping array and the ten genetic principal components. Genome-wide significance was *P <* 5 × 10^−8^. The covariate-adjusted comparison (Fig. 2d,e) re-ran the *raw*-embedding per-axis GWAS with BMI, height and imaging centre added as additional model covariates, holding all else fixed.

### 4.8 Heritability, intercepts and genetic correlation

SNP-heritability, LD-score intercepts and genetic correlations were estimated with LD-score regression ^11,18^. Summary statistics were harmonised with `munge_sumstats.py` to HapMap3 SNPs (∼1.17M variants), excluding the extended MHC, strand-ambiguous and low-MAF variants, and regressed against precomputed European LD scores. An axis was called significantly heritable at heritability *z >* 3. Bivariate (cross-trait) LD-score regression was used for all genetic correlations; because it is robust to sample overlap through the freely estimated cross-trait intercept, the same imaged sample could be correlated against both external and in-sample reference traits. Reference and disease GWAS are listed under Data availability.

### 4.9 Omnibus test, calibration and locus definition

The 20 orthogonal axes were combined into a single per-variant omnibus statistic, 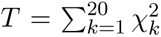, distributed as 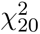 under independence of the axes. Independence was validated with MOSTest ^13^: the empirical between-axis *z*-score correlation matrix had off-diagonal entries in [−0.08, 0.05]. Calibration was assessed by quantile–quantile comparison of *T* across 14.7 M variants to the 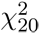 null; the bulk genomic-inflation analogue, computed after excluding genome-wide-significant variants, was *λ*_bulk_ = 1.04. Genome-wide-significant omnibus variants (*P <* 5 × 10^−8^) were merged into independent loci by grouping lead variants within 1 Mb, yielding 101 loci. A prespecified phenotype-level filter, applied independently of the association results, then removed loci driven by axis PC4 (the axis with the largest residual genetic correlation with body-fat percentage, *r_g_* = −0.24, *p <* 10^−3^) and loci in the MHC (chr6:25–35 Mb), leaving 84 confound-controlled cardiac-associated loci. Genes are reported as positional (nearest-gene) annotations of the lead variant unless colocalisation provided a specific candidate.

### 4.10 Robustness of the correction and discovery

We stress-tested the pipeline along five axes. (i) *Nuisance specification*: the residualisation was re-fit dropping BMI, dropping height, or retaining only imaging centre, and the full discovery re-run under each. (ii) *Cross-fitting*: as above. (iii) *Encoder seed*: the seed-2 crop encoder. (iv) *Dimensionality*: 10 and 40 retained axes. For each variant we recomputed the omnibus and re-derived the locus set, then quantified recovery as the fraction of the main 84-locus cardiac set whose lead variant was matched within 1 Mb by a genome-wide-significant lead in the alternative analysis, and separately confirmed recovery of every prespecified positional candidate.

### 4.11 Encoder-leakage and incremental-value controls

Because encoder pretraining and the GWAS drew on overlapping participants, we repeated the per-axis GWAS in 13,717 participants held out of encoder pretraining and computed lead-variant *β*-concordance against the full-sample analysis. To quantify value beyond expert phenotypes at equal power, we compared the learned-axis loci against a GWAS of 36 expert cardiac IDPs after randomly down-sampling the learned-axis GWAS to the expert-IDP sample size (≈33,000); a learned locus was counted as “learned-only” if no expert-IDP association had a genome-wide-significant lead within 1 Mb and in LD.

### 4.12 Colocalisation, disease and cardiac-trait genetic correlation

Statistical colocalisation between the learned-axis associations and GTEx v8 cis-eQTLs in cardiac and vascular tissues used the Bayesian coloc.abf framework ^19,20^ with default priors (*p*_1_ = *p*_2_ = 10^−4^, *p*_12_ = 10^−5^) over the cis-window around each lead variant; a shared causal variant was called at posterior PP.H4 *>* 0.5. Genetic correlation with coronary artery disease was estimated by cross-trait LD-score regression against two external GWAS: Aragam et al. ^14^ (UK-Biobank-inclusive) and Nikpay et al. ^15^ (CARDIoGRAMplusC4D, UK-Biobank-free, chosen to exclude sample overlap). Genetic correlation with a prespecified cardiac panel (atrial fibrillation, the electrocardiographic PR interval and heart failure) was estimated for all 20 axes; per-trait significance was assessed at *p <* 0.05*/*20 and panel-wide significance at *p <* 0.05*/*60.

### 4.13 Clinical outcomes

We assessed the prognostic value of the corrected representation with time-to-event analysis of four incident outcomes: atrial fibrillation, heart failure, coronary artery disease and cardiomyopathy. Cases were defined from the UK Biobank first-occurrence fields (which combine hospital-inpatient, death-register and primary-care linkage) as the first ICD-10 diagnosis recorded *after* the imaging visit; participants with the corresponding diagnosis on or before the imaging visit were excluded as prevalent. Follow-up ran from the imaging visit to the first event, death, or the end of record linkage, whichever occurred first. For each outcome we fitted three nested Cox proportional-hazards models (lifelines) on a common complete-case sample: age and sex; those covariates plus the expert cardiac-MRI imaging-derived phenotypes (left-ventricular volumes, ejection fraction and mass); and those plus the 20 corrected embedding axes. All continuous predictors were standardised, with standardisation parameters estimated on the training data only and applied unchanged to held-out data. Discrimination was quantified as Harrell’s concordance index averaged over five cross-validation folds that were disjoint at the participant level (no participant contributed to more than one fold), with every predictor transformed using training-fold parameters to prevent leakage. Incremental value was tested with a likelihood-ratio test comparing the expert-CMR and expert-CMR-plus-embedding models on the full complete-case data; we report this full-data *p* alongside the cross-validated C-index, which are complementary (the C-index estimates out-of-sample discrimination and the likelihood-ratio test the in-sample nested improvement). No predictor selection or penalisation was applied, the models carry no tuned hyperparameters, and the proportional-hazards assumption was examined with scaled Schoenfeld residuals. The complete-case analysis samples comprised 28,917 participants and 571 events for atrial fibrillation, 29,580 and 205 for heart failure, 29,652 and 756 for coronary artery disease, and 29,605 and 53 for cardiomyopathy; cardiomyopathy was underpowered and is reported without emphasis.

### 4.14 Anatomical interpretability

To characterise where within the imaged heart a corrected axis varies, we contrasted participants at the low and high extremes of the axis (bottom versus top decile of the axis score). For each imaging plane (2-, 3- and 4-chamber) and cardiac phase (end-diastole, end-systole) we formed the per-pixel mean cine frame within each extreme group on the motion-cropped field of view, and computed a per-pixel Cohen’s *d* effect-size map, *d* = (*x̅*_high_ − *x̅*_low_)*/s*_pooled_, where *s*_pooled_ is the pooled within-group standard deviation. Structure confined to discrete cardiac walls, rather than a diffuse whole-frame shift, indicates that the axis varies with specific cardiac structures within the field of view; because only cardiac images were analysed, this does not test specificity relative to other organs.

### 4.15 Use of large language models

Large language model-based assistants were used to support manuscript drafting, editing and code refactoring. All study design, analyses, figures, results and their interpretation were performed and verified by the authors, who take full responsibility for the content of this work.

### 4.16 Software, seeds and reproducibility

Association testing used REGENIE v4; genotype handling used PLINK 2.0; heritability and genetic correlation used LD-score regression (`ldsc`, Python) with `munge_sumstats.py`; colocalisation used the `coloc` R package; survival analysis used `lifelines` (Python). Representation learning used PyTorch, and the decodability probes used `scikit-learn` (ridge regression and random-Fourier-feature approximation). Data wrangling used `pandas/numpy` (Python) and `data.table` (R); figures were produced in R with `ggplot2` and `patchwork`. All stochastic steps used a fixed random seed (42) unless a second seed is explicitly reported. Key thresholds were: genome-wide significance *P <* 5×10^−8^; heritability call *z >* 3; colocalisation PP.H4 *>* 0.5; independent-locus merge distance 1 Mb; MHC exclusion chr6:25–35 Mb. The statistical-genetics pipeline and the figure-generation code are available at the repository listed under Code availability; the encoder-training and representation-processing scripts, and aggregate per-axis summary statistics sufficient to regenerate every figure, are available from the corresponding author on request.

## Ethics approval and consent to participate

UK Biobank has ethical approval from the North West Multi-centre Research Ethics Committee (REC reference 11/NW/0382), and all participants provided written informed consent at recruitment. This research was conducted under UK Biobank approved application 65439. No additional ethical approval was required for the secondary analyses reported here.

## Data availability

Individual-level UK Biobank data are available to approved researchers via the UK Biobank Access Management System (https://www.ukbiobank.ac.uk) under application 65439. All external datasets used are publicly available: the CARDIoGRAMplusC4D 1000 Genomes-based coronary artery disease meta-analysis (http://www.cardiogramplusc4d.org; GWAS Catalog GCST003116); the Aragam et al. coronary artery disease genome-wide association study (GWAS Catalog GCST90132314); GTEx v8 cis-eQTL summary statistics (https://www.gtexportal.org); and the reference GWAS used for genetic-correlation analyses (atrial fibrillation, PR interval, heart failure, and anthropometric and skeletal traits) obtained through LD Hub and the GWAS Catalog. Aggregate per-axis summary statistics and the figure-source tables are available from the corresponding author on request.

## Code availability

All analysis code is publicly available at https://github.com/Devanshpandey/ssl-imaging-genetics and will be archived on Zenodo upon publication. The repository contains code only; no UK Biobank or individual-level data are included.

## Acknowledgements

Computing support for the project was provided through the Director’s Discretionary Fund at the Texas Advanced Computing Center (TACC) and used the Lonestar6 system. V.M.N. was supported by a grant for human brain evolution through the Allen Discovery Center program, a Paul G. Allen Frontiers Group advised program of the Paul G. Allen Family Foundation, and by a Good Systems Fellowship for Ethical AI at The University of Texas at Austin. D.P. was supported by an Outstanding Graduate Research Fellowship from the University of Texas at Austin Graduate School. This research was conducted using the UK Biobank Resource under application 65439.

## Author contributions

D.P. designed the study, implemented the pipeline, performed all analyses, and wrote the manuscript. V.M.N. supervised the study, provided funding and computational resources, and revised the manuscript. Both authors approved the final manuscript.

## Supplementary Table

**Supplementary Table S1.**
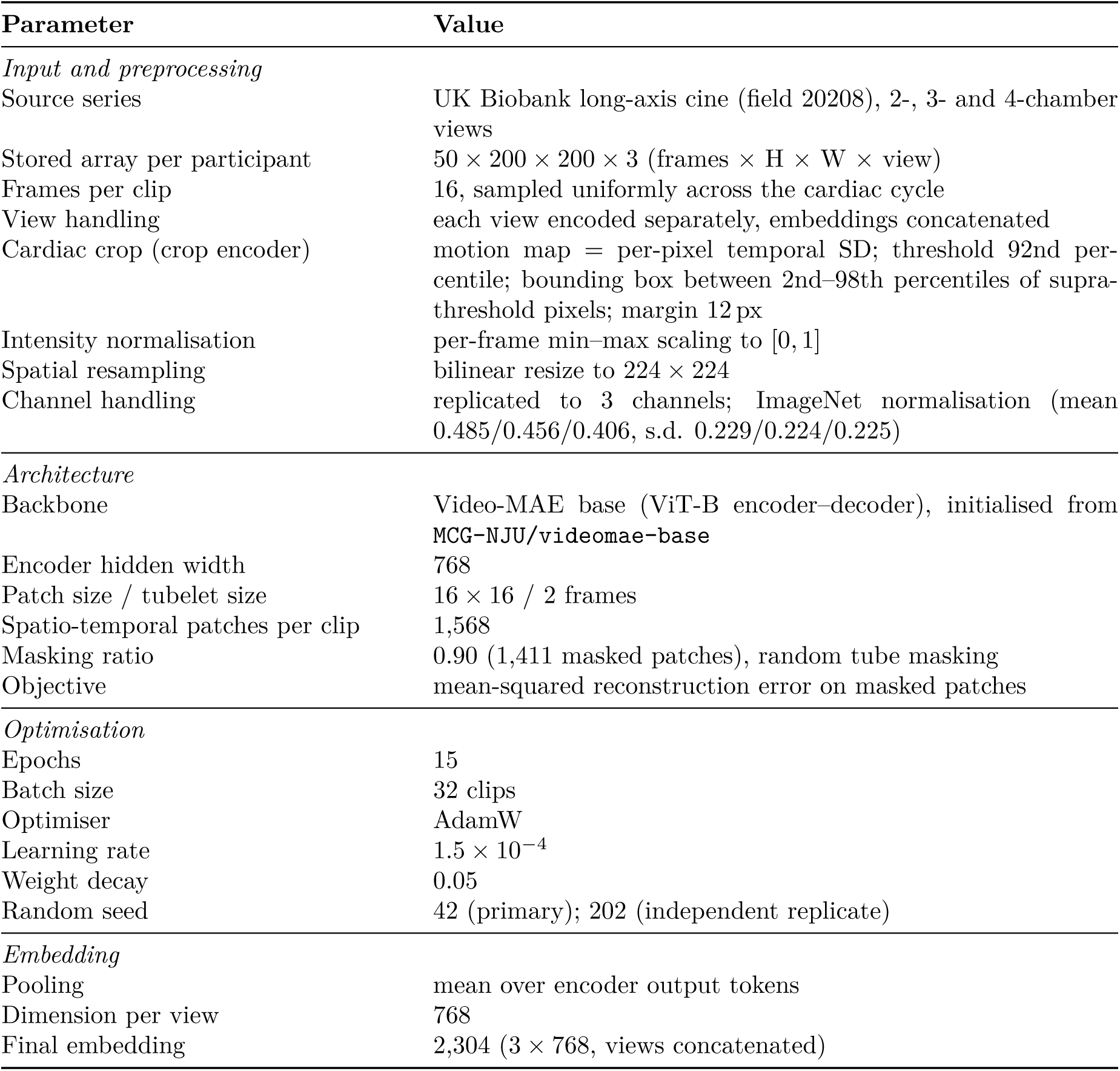
Video-MAE preprocessing and training parameters. Values are those used for the encoders reported in this study. The base (full-field-of-view) and motion-crop encoders share all settings except the cardiac crop.

## Supplementary Figures

**Figure S1:**
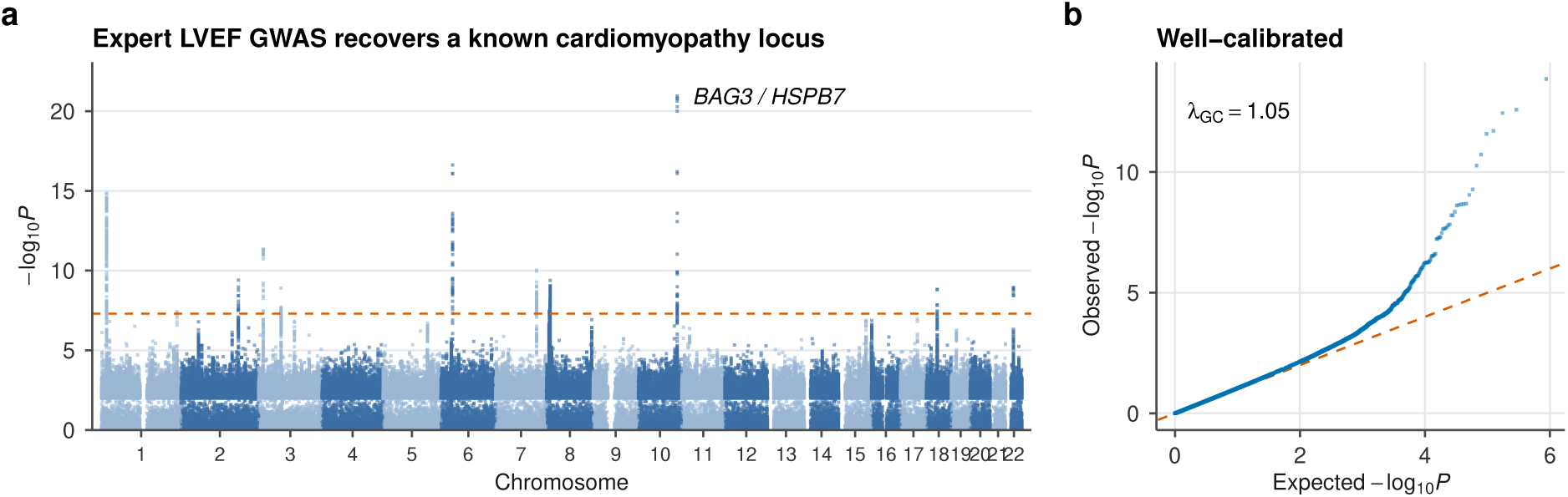
Positive control: the pipeline recovers known cardiac biology and is well calibrated. **(a)** Manhattan plot of an expert-LVEF GWAS in the imaged cohort, recovering *BAG3* /*HSPB7*. **(b)** Quantile–quantile plot; *λ*_GC_ = 1.05 indicates well-calibrated statistics, with an upward tail consistent with polygenic signal.

**Figure S2:**
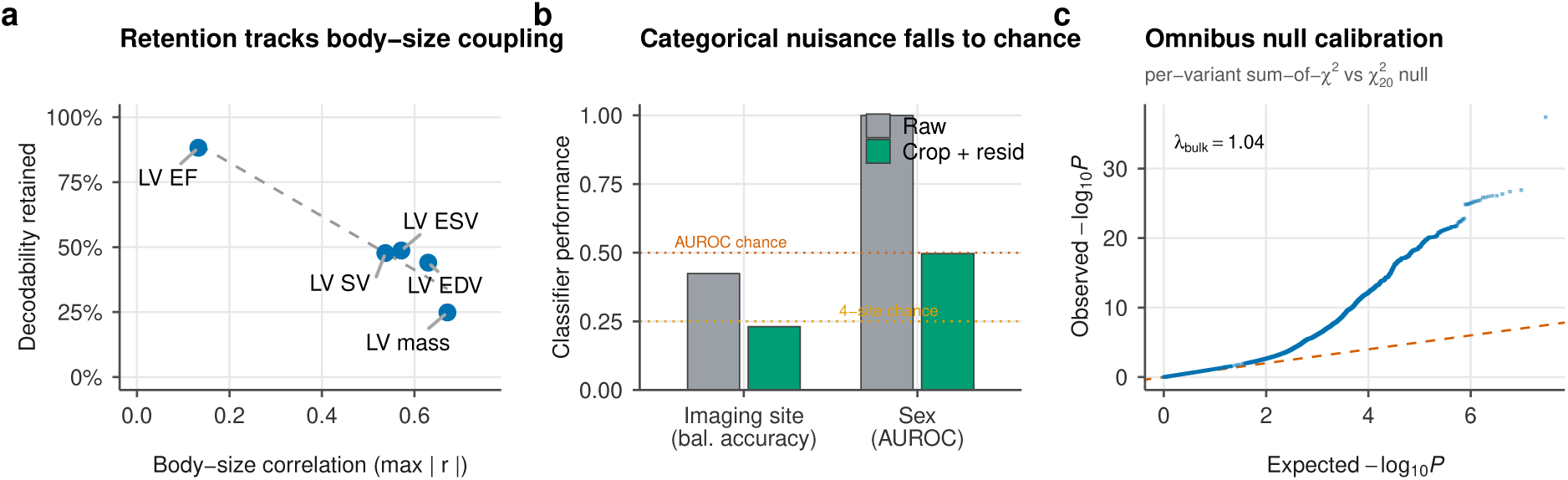
Additional controls on the correction. **(a)** Fraction of cardiac-phenotype decodability retained after correction versus the phenotype’s body-size correlation (max |*r*| with BMI or height); retention falls monotonically, so the correction preferentially attenuates the body-size-coupled component. **(b)** Categorical nuisance decoding with conventional classifier metrics: sex (AUROC 1.00 → 0.50) and imaging site (balanced accuracy 0.42 → 0.23, four-site chance 0.25) both fall to chance after correction. **(c)** Omnibus null calibration: quantile–quantile plot of the per-variant sum-of-*χ*^2^ statistic against its 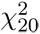 null; the bulk inflation analogue is *λ*_bulk_ = 1.04 and the upper-tail departure is consistent with polygenic signal.

**Figure S3:**
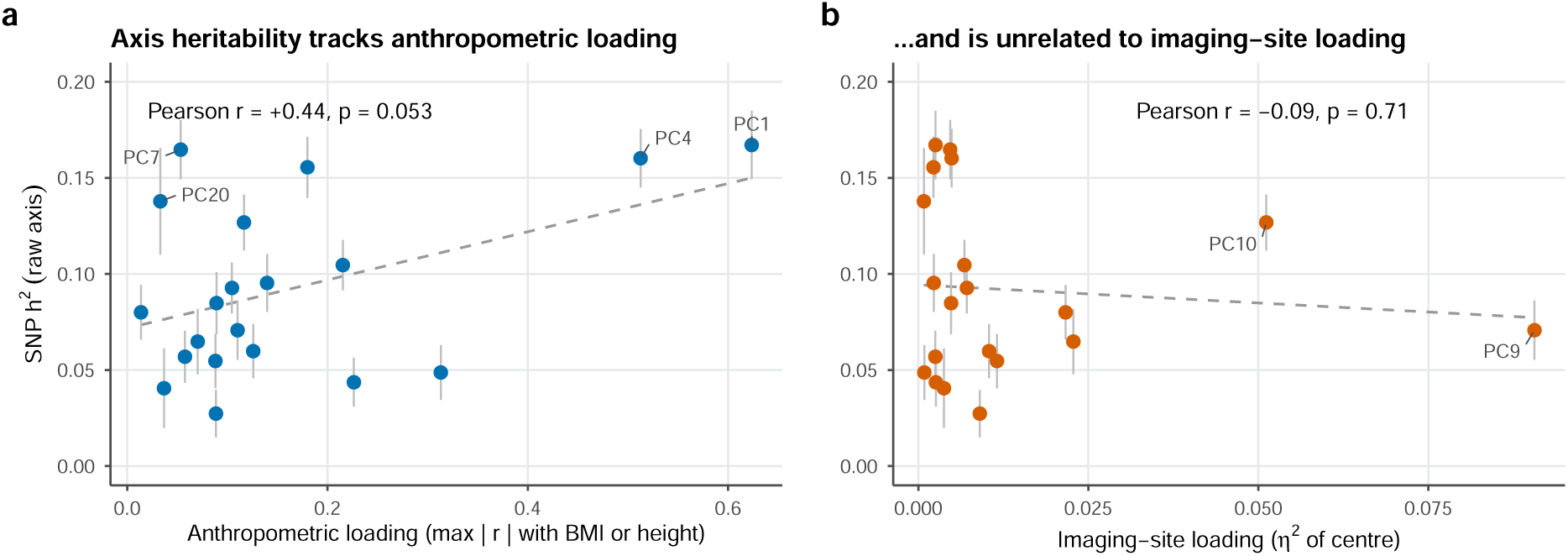
Relationship of axis heritability to anthropometric and imaging-site loading. Per-axis SNP-heritability (LD-score regression; error bars, standard error) for the 20 raw latent axes plotted against **(a)** anthropometric loading (maximum |*r*| with BMI or height) and **(b)** imaging-site loading (*η*^2^ of assessment centre), computed in the 54,745 participants with complete covariates. Heritability shows a positive association with anthropometric loading (Pearson *r* = 0.44, *p* = 0.053); no association is observed with site loading (*r* = −0.09, *p* = 0.71). Imaging site contributes phenotypic but not genetic variance, diluting rather than abolishing SNP-heritability, so a site-decodable representation can nonetheless be heritable through its anthropometric components. Dashed lines, ordinary least-squares fits.

**Figure S4:**
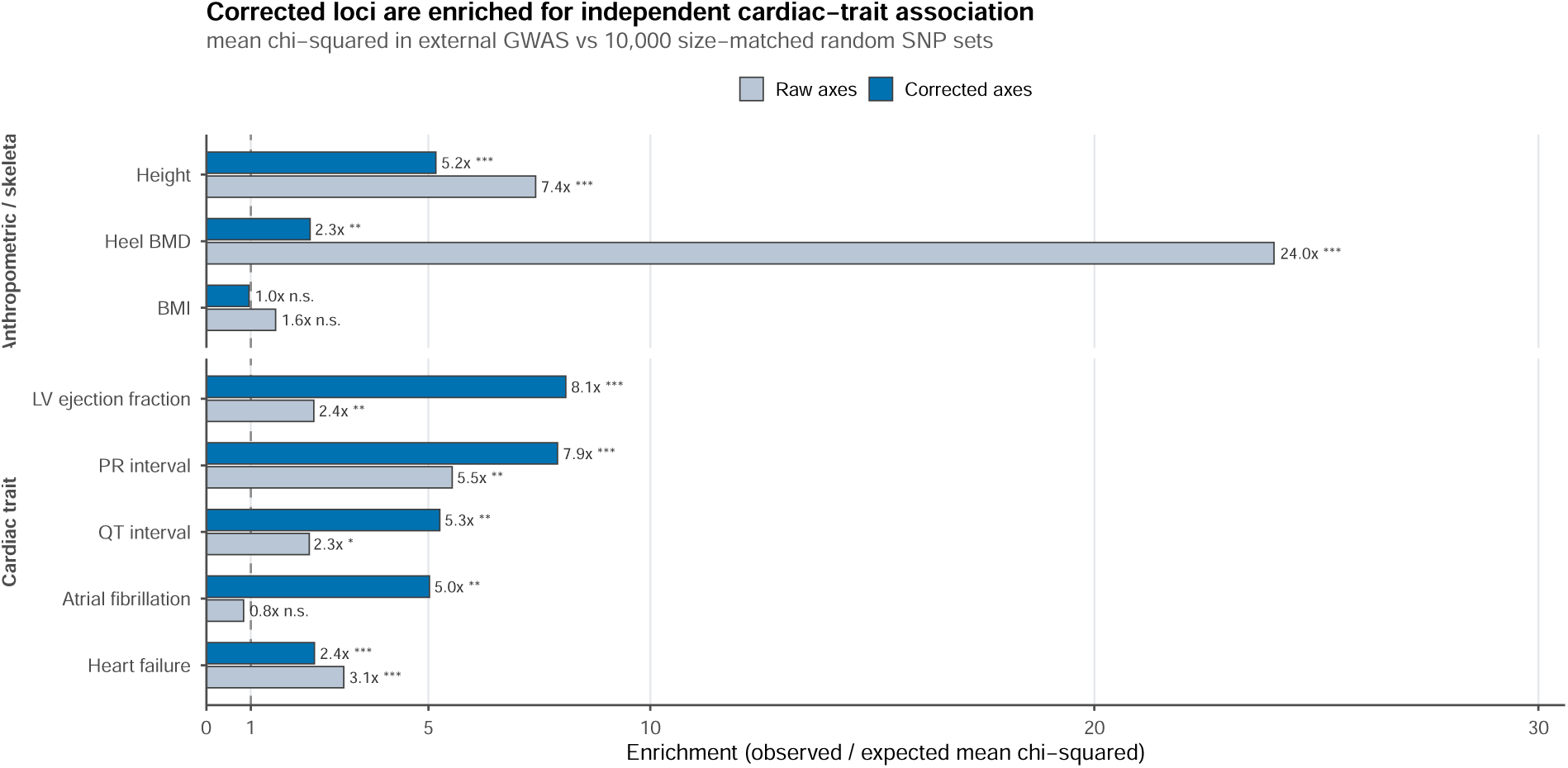
Correction shifts locus-level enrichment from skeletal towards cardiac traits. Enrichment of lead variants for association in independent external GWAS, computed as the observed mean *χ*^2^ of the lead-variant set divided by the mean of 10,000 size-matched random SNP sets drawn from the same summary statistics; the dashed line marks no enrichment. Blue, the 84 confound-controlled cardiac loci; grey, loci from the raw (uncorrected) axes. Correction increases enrichment for cardiac-function and conduction traits (LV ejection fraction, QT interval, atrial fibrillation) and reduces skeletal enrichment (heel bone-mineral density 24.0×→ 2.3×), with BMI enrichment null after correction. Residual height enrichment is consistent with cardiac allometry. Permutation *p*: ∗ *<* 0.05, ∗∗ *<* 0.01, ∗ ∗ ∗ *<* 0.001. Overlap with each reference panel varies (20–64 of 84 lead variants present), because the HapMap3-restricted reference summary statistics differ in coverage.

